# Dysphagic disorder in a cohort of COVID-19 patients: evaluation and evolution

**DOI:** 10.1101/2021.06.20.21258947

**Authors:** Andrea Glotta, Anna Galli, Maira Biggiogero, Giovanni Bona, Andrea Saporito, Romano Mauri, Samuele Ceruti

## Abstract

**Background:** COVID-19 is a multisystem disease complicated by respiratory failure requiring sustanined mechanical ventilation (MV). Prolongued oro-tracheal intubation is associated to an increased risk of dysphagia and bronchial aspiration. Purpose of this study was to investigate swallowing disorders in critically ill COVID-19 patients.

**Methods:** This was a retrospective study analysing a consecutive cohort of COVID-19 patients admitted to the Intensive Care Unit (ICU) of our Hospital. Data concerning dysphagia were collected according to the *Gugging Swallowing Screen (GUSS)* and related to demographic characteristics, clinical data, ICU Length-Of-Stay (LOS) and MV parameters.

**Results:** From March 2 to April 30 2020, 31 consecutive critically ill COVID-19 patients admitted to ICU were evaluated by speech and language therapists (SLT). Twenty-five of them were on MV (61% through endotracheal tube and 19% through tracheostomy); median MV lenght was 11 days. Seventeen (54.8%) patients presented dysphagia; a correlation was found between first GUSS severity stratification and MV days (p < 0.001), ICU LOS (p < 0.001), age (p = 0.03) and tracheostomy (p = 0.042). No other correlations were found. At 16 days, 90% of patients had fully recovered; a significant improvement was registered especially during the first week (p < 0.001).

**Conclusion:** Compared to non-COVID-19 patiens, a higher rate of dysphagia was reported in COVID-19 patients, with a more rapid and complete recovery. A systematic early SLT evaluation of COVID-19 patients on MV may thus be useful to prevent dysphagia-related complications.

## INTRODUCTION

CoronaVirus Disease-19 (COVID-19) is a viral pneumonia caused by the novel Severe Acute Respiratory Syndrome Coronavirus 2 (SARS-CoV-2). Since December 2019 the disease spread quickly around the world, currently affecting more than 115’000’000 patients worldwide ^1^. Invasive mechanical ventilation (MV) is a support therapy often required by critically ill COVID-19 patients affected by acute lung injury ^2,3^. Since this complication is usually associated with prolonged MV, in some case mandating tracheostomy is required ^4^.

A known complication of prolonged MV and tracheostomy is pharyngeal muscles dysfuntion, associated with dysphagia ^5^ and swallowing disorders ^6^. Rather than a disease, dysphagia is a symptom, which is often associated to several complications ^7^ that can potentially lead to a significant increase of both intensive care unit (ICU) length-of-stay LOS and in overall in-hospital mortality ^8^. Among these, aspiration pneumonia incidence has been reported to be 11 times greater in dysphagic compared to non-dysphagic ICU patients ^9^.

Clinical studies investigating the association between the duration of MV and dysphagia reports conflicting results ^5,10–13^. Laryea et al ^13^, Ajeman et al ^10^ and Brodsky et al ^6,11,14^ found an association between dysphagia and prolonged orotracheal intubation, while two systematic reviews by Skoretz et al ^5,12^ concluded that further studies are needed in order to confirm a definitive correlation. Only one review summarizes the available information on possible mechanisms of postintubation dysphagia in COVID-19 patients, while very few data concerning the relationship between dysphagia and prolonged orotracheal intubation have been reported in these patients ^15^.

Aim of this study was to investigate the prevalence of dysphagia in critically ill COVID-19 patients, describing the features of the swallowing disorders detected and the associated risk factors.

## METHODS

The Canton Ticino was one of the Swiss Regions worstly affected by the COVID-19 pandemic ^16^; more than 1613 patients were admitted at our Hospital for this disease and more than 100 patients were admitted to the ICU. We conducted a retrospective observational study on a consecutive cohort of patients affected by COVID-19 requiring MV, admitted to our ICU since March 2^nd^ to April 30^th^ 2020. After local Ethical Committee approval (Comitato Etico Cantonale, ref. n. CE_TI_3692), written informed consents from each patient have been obtained before data collection. All patients were systematically evaluated by the speech and language therapists (SLT) team. Patients who died before the first evaluation and patients not on MV were excluded from the analysis. All collected data were reported on an electronic database.

### SLT evaluation

#### Dysphagia assessment evaluation

In order to minimize the risk of contagion, SLT evaluations consisted of a clinical evaluation only. Among different options availale to assess and quantify dysphagia, like the Toronto bedside swallowing screen test ^17^, the Massey bedside swallowing screen ^18^, the Daniel’s test, the GUSS-ICU ^19^, the Disphagia Outcome and Severity Scale ^20^ and the Penetration-Aspiration Scale ^21^, the Gugging Swallowing Screen (GUSS) ^22^ was chosen. This tool resulted to be more precise compared to the other ones, as it provides a dysphagia severity score that takes into account food texture, correlating it with the severity of the dysfuntion.

#### Evaluation test and timing

SLT evaluation was consisted in a preliminary assessment followed by a direct swallowing test ^22^. GUSS scoring system implies 4 categories of severity, well describing the functional degree of dysphagia: severe dysphagia (0-9 points), moderate dysphagia (10-14 points), mild dysphagia (15-19 points) and absence of dysphagia (20 points). Starting from the first evaluation at day 0, GUSS was performed daily; after discharge from the ICU, patients were daily re-evaluated according to GUSS scale until hospital discharge or complete recovery.

#### SLT treatment

The evaluation was completed following the Logemann protocol procedures ^23^. Based on the global evaluation (GUSS and Logemann protocol procedure), a rehabilitation programme was structured. Its aims were to reduce the oral and pharyngeal sensitivity deficit, the delay of swallowing reflex start ^21,24–26^, and the glottic closure deficit ^27^ as well as to improve laryngeal elevation motor skills ^28,29^.

### Data recording

For each patient, SLT assessments were performed at day 0, day 7, day 14, day 21 and day 28, both in ICU and after transfer to the Internal Medicine ward. Additional demographic factors like age, body-mass index (BMI) and comorbidities like chronic-obstructive pulmonary disease (COPD), obstructive sleep apnea syndrome, diabetes, hypertension and ischemic heart disease (IHD) were registered and reported. Evaluation of ICU severity scores NEMS (nine equivalents of nursing manpower use score), SAPS (simplified acute physiology score) and SOFA (sequential organ failure assessment) at ICU admission were calculated for every patient included.MV parameters, days of MV, ICU LOS, number of pronation manouvers performed and positive end-expiratory pressure (PEEP) values were also recorded for all cases. Finally, complications like ventilation-associated pneumonia (VAP), the need of continuous renal replacement therapy (CRRT) and venous-thromboembolism (VTE) were also registered and analyzed.

### Statistical analysis

Descriptive statistics of frequency was performed. Data were reported as number (percentage). Data distribution was reported as mean (SD) if normally distributed, otherwise as median (IQR). Data distribution was verified by Kolmogorov-Smirnov test. The relationship between GUSS values and continuous variables was analized by linear regression. Differences between continuous variables were studied by t-test; categorical data differences were carried out by Chi-square analysis. All statistical tests were two-tailed; significance level was established to be < 0.05. Statistical data analysis was performed using the SPSS 26.0 package (SPSS Inc, USA).

## RESULTS

### Demographics

During the study period, 42 consecutive critically ill COVID-19 patients were admitted to our ICU; 11 of them died and were consequently excluded from the study. Thirty-one patients were systematically evaluated by the SLT team. Mean age was 61 years (SD 12 yrs) and 25 patients (80.6%) were male, with a mean BMI of 29 Kg/m^2^ (SD 4.8). At admission, median NEMS score was 34 (18 – 39), mean SAPS score was 43 (SD 18) and mean SOFA score 6 (SD 2.8) (Table 1).

**TABLE 1:** Demographics.

|  | Characteristics | Enrolled (n = 31) |
| --- | --- | --- |
| Demographics | Age | 61 (29 – 76, 12) |
|  | Male | 25 (80.6%) |
|  | BMI | 29 (20 – 41, 4.8) |
| Comorbidities | COPD | 1 (3.2%) |
|  | OSA | 4 (12.9%) |
|  | Diabetes | 10 (32.3%) |
|  | Hypertension | 13 (41.9%) |
|  | IHD | 4 (12.9%) |
| Severity score at ICU admission | NEMS | 34 (18 – 39) |
|  | SAPS | 43 (13 – 94, 18) |
|  | SOFA | 6 (0 – 11, 2.8) |
| Complication | VAP | 4 (12.9%) |
|  | CRRT | 4 (12.9%) |
|  | VTE | 4 (12.9%) |
| MV parameters | Patients on MV | 25 (80.64%) |
|  | • Endotracheal tube | 19 (76%) |
|  | • Tracheostomy | 6 (24%) |
|  | Pronation maneuvers | 3 (0 – 8, 2.6) |
|  | PEEP | 10 (19 – 15) |
| Outcomes | ICU LOS | 13 (11 – 19) |
|  | MV Days | 11 (7.5 – 16) |

Demographic characteristics at ICU admission. Data are presented as means (min-max, SD) or medians (IQR) for non-normally distributed variables.

Of the 31 evaluated patients, 25 (80.64%) were on invasive MV; 19 (76%) were ventilated via an endotracheal tube and 6 (24%) underwent tracheostomy at some point, due to a prolonged MV. Median ICU LOS was 13 days (9-11) and median MV days were 11 (7.5–16). Patients ventilated via an endotracheal tube presented a mean MV days of 11.5 (SD 7.7) while patients undergoing tracheostomy presented a mean MV days of 20.5 (SD 11.6). In order to verify if tracheostomy has been determined by the previous MV days, a two-tailed T-test has been performed. No significant difference was found between MV days in patients ventilated via an endotracheal tube and MV days in patients ventilated via tracheostomy (t-test 1.780, p = 0.122).

Ten (32.25%) patients presented complications, such as VAP (4–12.9%), acute kidney injury requiring CRRT (4–12.9 %) and VTE (4–12.9%); 2 patients showed more than one of these complications. At the first SLT evaluation, median GUSS was 19 (12-20), with a mean of 15. Fourteen (45.2%) patients presented no dysphagia (GUSS equal to 20), 5 (16.1%) had the criteria for mild dysphagia (GUSS 15-19), 6 (19.4%) for moderate dysphagia (GUSS 10-14) and 6 (19.4%) for severe dysphagia (GUSS less than 9).

### GUSS scores correlations

On day 0, GUSS values showed an inverse correlation with MV duration (r^2^ = 0.616, p < 0.001), ICU LOS (r^2^ = 0.558, p < 0.001), age (r^2^ = 0.392, p = 0.03) and tracheostomy (p = 0.04) (Figure 1, Table 2). No correlation was found between GUSS values and other variables, such as sex (p = 0.407), BMI (p = 0.67), NEMS at admission (p = 0.77), SAPS at admission (p = 0.52), SOFA at admission (p = 0.45), number of pronation maneuvers performed (p = 0.98) and initial PEEP (p = 0.35). Similarly, no correlation was found between GUSS values registred at day 0 and comorbidities (COPD p = 0.06, OSA p = 0.6, diabetes p = 0.20, arterial hypertension p = 0.48, ischemic heart disease p = 0.72) or incidence of clinical complications (VAP p = 0.14, CRRT p = 0.72, VTE p = 0.59) (Table 2).

**TABLE 2:** SLT correlations at day 0.

|  | Variable | Correlation | Chi-square | P value |
| --- | --- | --- | --- | --- |
| Correlation between<br>GUSS at day 0 and | Age | 0.39 | - | 0.03* |
|  | Sex | - | 8.27 | 0.41 |
|  | BMI | 0.08 | - | 0.67 |
|  | COPD | - | 14.98 | 0.06 |
|  | OSA | - | 6.42 | 0.60 |
|  | Diabetes | - | 10.95 | 0.20 |
|  | Hypertension | - | 7.53 | 0.48 |
|  | IHD | - | 5.36 | 0.72 |
|  | NEMS score | 0.05 | - | 0.77 |
|  | SAPS Score | 0.12 | - | 0.52 |
|  | SOFA Score | 0.14 | - | 0.45 |
|  | MV days | 0.78 | - | < 0.001* |
|  | ICU LOS | 0.78 | - | < 0.001* |
|  | VAP | - | 12.35 | 0.14 |
|  | CRRT | - | 5.36 | 0.72 |
|  | VTE | - | 13.99 | 0.59 |
|  | Pronation | 0.004 | - | 0.98 |
|  | PEEP | 0.17 | - | 0.35 |
|  | Tracheostomy | - | 16.05 | 0.04* |

Correlation between GUSS evaluation at day 0 and demographics characteristics. Significant correlation with p-value < 0.05

**Fig. 1.**
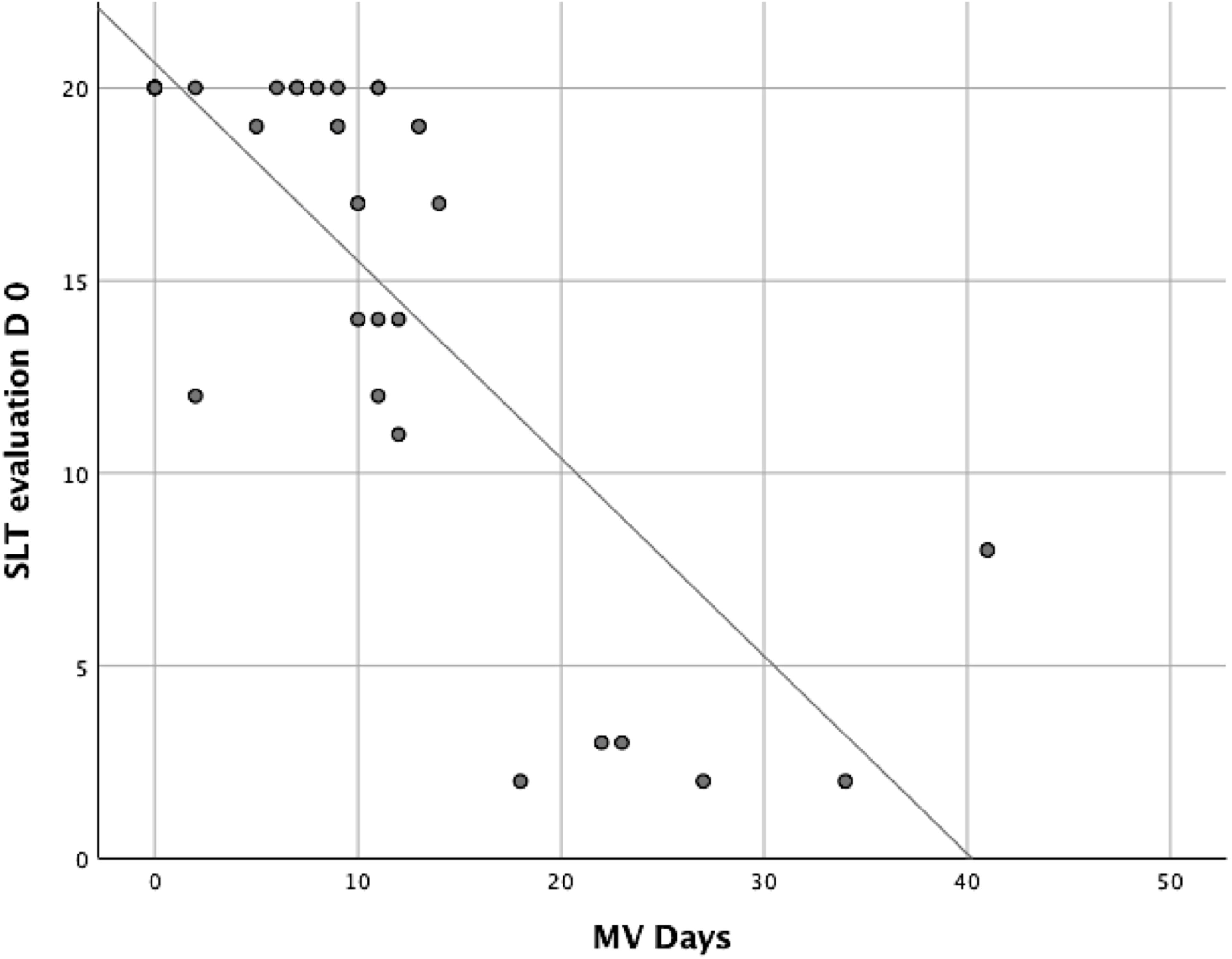
Linear regression between first SLT evaluation (day 0) and of MV days (r^2^ = 0.616).

### GUSS scores evolution

At day 7 since first SLT assessment, all patients’ GUSS score improved; GUSS score stratification identified 2 (6.5%) patients with severe dysphagia, 2 (6.5%) with moderate dysphagia, 4 (12.9%) with mild dysphagia and 23 (74.2%) without dysphagia. This score evolution resulted statistically different (p < 0.001) from the baseline at day 0. Severe, moderate, mild and no dysphagia evolution showed a prevalence of 0%, 3.2%, 9.7%, 87.1% respectively at day 14 (p = 0.01), a prevalence of 0%, 3.2%, 6.5%, 90.3% respectively at day 21 and of 0%, 3.2%, 0, 96.8% respectively at day 28 (Figure 2). According to GUSS evaluation, all patients showed a progressive improvement throughout the days; at day 12 severe forms of dysphagia were no longer registered; moreover, at day 16 90% of patients fully recovered from swallowing disorder, with a GUSS of 20 (Figure 3, e-Figure 2).

**Fig. 2.**
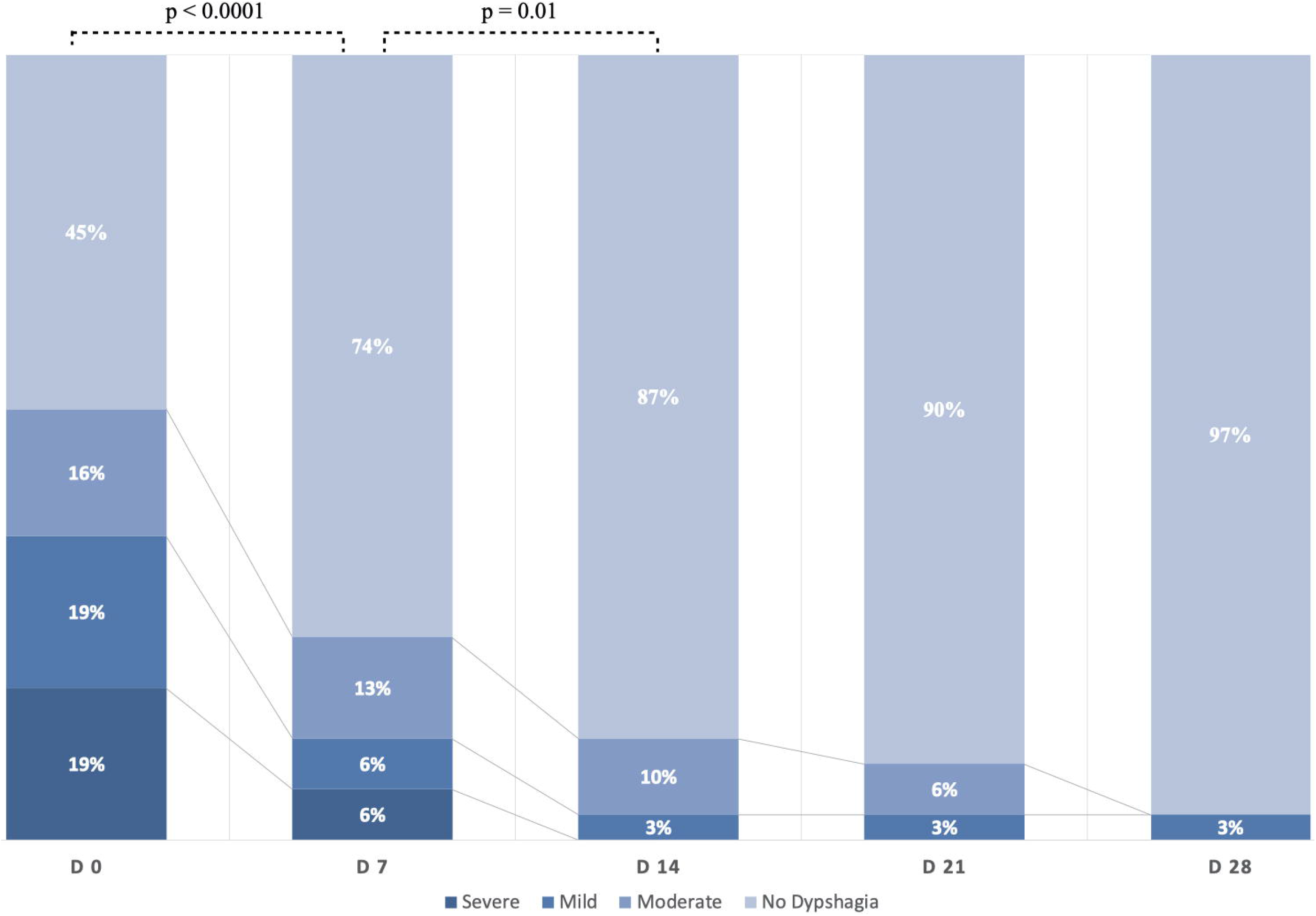
Temporal evolution of dysphagia according to GUSS value, stratified into four semi-quantitative groups at different evaluation timepoints. GUSS evolution between day 0 and 7 and between day 7 and 14 were statistically significant (p < 0.0001 and p = 0.01 respectively).

**Fig. 3.**
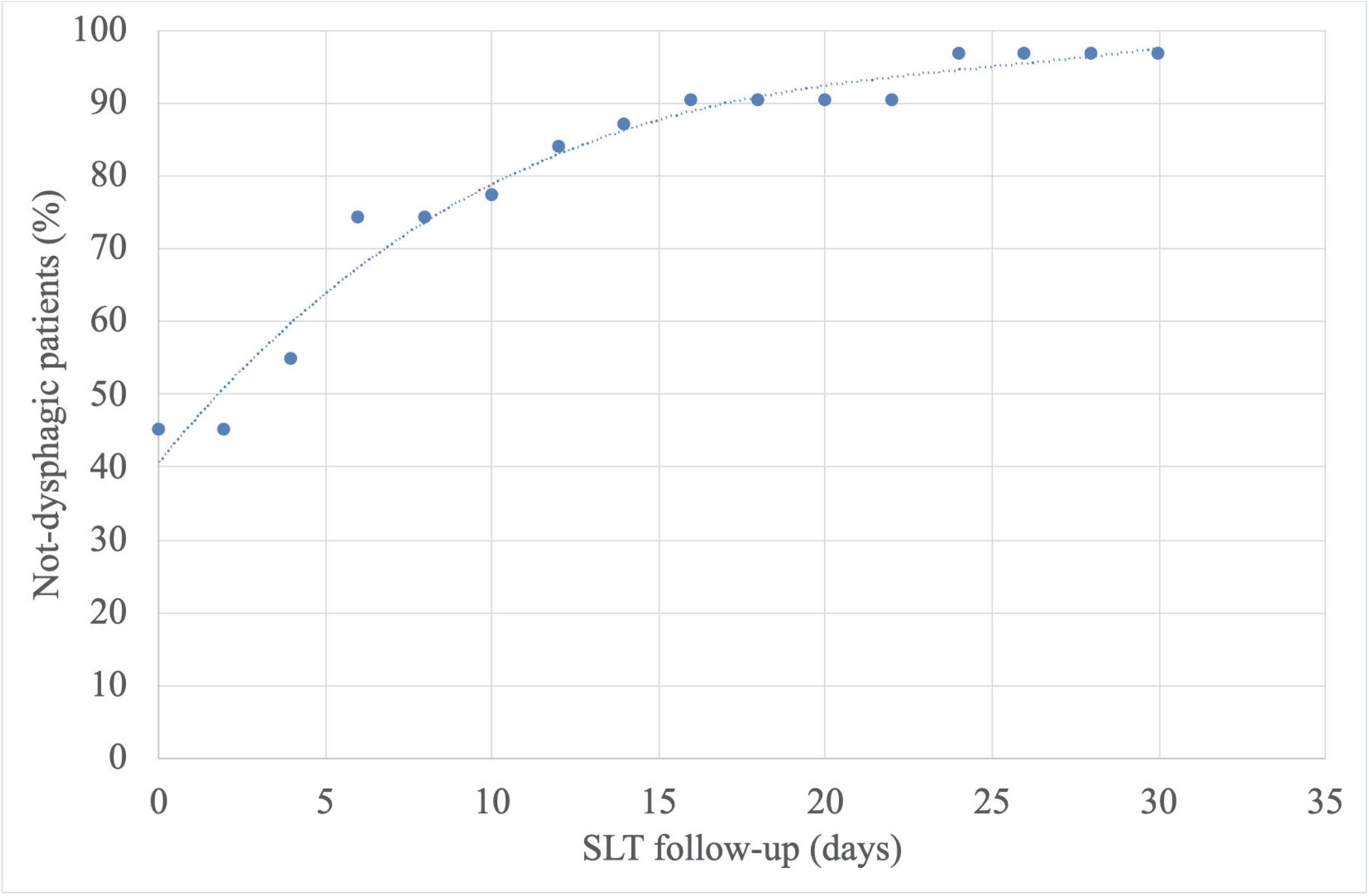
Improvement in patients swallowing disorder (GUSS = 20, no dysphagia) from first SLT evaluation at day 0 up to day 31 of follow up.

At the end of the study, 16 (52%) patients were discharged at home, 12 (38%) were transferred in post-acute rehabilitation institutions, 1 (3.3%) was transferred to other hospitals, 1 (3.3%) died and 1 (3.3%) was still hospitalized. At hospital discharge 7 (22.5%) patients still presented a mild degree of dysphagia (GUSS between 15 and 19); all of these patients were transferred to rehabilitation facilities.

## DISCUSSION

Critically ill COVID-19 patients usually require a prolonged MV and a long ICU stay, conditions associated to a high mortality rate ^30,31^. Moreover, even in case of survival, important morbidities like dysphagia can occur, with a possible severe impact on quality of life ^32,33^. The aim of the study was the assessment of both prevalence and degree of dysphagia in critically ill COVID-19 patients, with the intention to give a valuable insight into this condition, which may affect the clinical outcome of these patients.

Following a specific ’care map’ procedure, patients included in this study presented a shorter MV and lower PEEP requirements, compared to other groups reported in literature ^34,35^. Despite this, our cohort showed a relevant prevalence of swallowing disorder, with more than half of patients (55%) with a certain degree of dysphagia at the first evaluation. In patients ventilated for more than 10 days, prevalence of dysphagia was even higher, up to 95.5%. According to literature, in non-COVID-19 patients ^7,36,37^, severe dysphagia seems to be strictly correlated with MV length and ICU LOS. In our study, we reported for the first time data on dysphagia in COVID-19 patients, who seem to be burdened by a higher incidence of dysphagia already at a very early stage, when compared to other patients, as seen in the studies of Kim et al ^7^, Oliveira et al ^36^ and Yang et al ^37^. This may be explained by the strong inflammatory response induced by SARS-CoV-2 and by its consequent systemic effects, potentially triggering a severe generalized neuromuscular impairment also involving pharyngeal muscles.

This hypothesis is further supported by the absence of a correlation between the ICU admission severity scores (SAPS, SOFA, NEMS) and the dysphagic disorders degree registered at the first SLT evaluation. This data would underscore the importance of all MV-related features as predictives for dysphagia incidence ^38,39^; at the same time, MV features could represent the severity of COVID-19 disease. Patients’ age was the only element identifiable at ICU admission that acted as predictive factor for a stronger and faster loss of muscle activities.

Even if the prevalence of dysphagia was high, the functional recovery appeared to be relatively fast, with a complete regresion of all severe cases of dysphagia after 12 days since the beginning of SLT assessment. The recovery time reported in non-COVID-19 patients is longer ^14,40^. These data, if confirmed by further appropriately designed studies, may suggest that SLT plays a role in COVID-19 patients global management; this is further suggested by data showing a possible shortening of the recovery time and an increase in the probability of swallowing disorders regression when SLT is implemented ^41,42^.

This study has some limitations. Firstly, it is a single center observational study; further studies will be necessary to confirm these preliminary data. Secondly, due to the COVID-19 emergency situation, it was not possible to determine a control group; we decided therefore to compare our data with the available literature. Thirdly, it was not possible to compare GUSS scale between different SLT teams, potentially leading to the presence of bias due to a single specialist evaluation. Finally, it was not possible to quantify the SLT’s work and its impact in patients’ dysphagia management.

## CONCLUSION

Critically ill COVID-19 patients presented a higher incidence of swallowing disorder than reported in non-COVID-19 patients; severe inflammatory dysregulation could explain the increased rate of pharyngeal neuromuscular impairment. Despite high dysphagia prevalence and severity, a short recovery period was reported. Finally, SLT could play a relevant role in critically ill COVID-19 patients’ multidisciplinary management.

## Data Availability

All data are available under reasonable request

## Authors’ contributions

AG, AG, GT and CM contributed to study design and data collection. AG, AG, MB, GT, CM, SC contributed to data analysis and interpretation. AG, AG, MB, AS, RM, XC, SC contibuted to write the manuscript.

### Conflicts of interest

The Authors declare no conflict of interest linked to the submitted work

### Funding

No funding has been required for this project

## Notes

### Competing Interest Statement

The authors have declared no competing interest.

### Author Declarations

Ethics committees: Comitato Etico Cantonale, Via Orico 4, 6500 - Bellinzona Ethical approval was given, with the internal referral number CE_TI_3692

